# Study protocol: Longitudinal observational study on frailty and mental health

**DOI:** 10.64898/2026.04.01.26349941

**Authors:** Petra Mikolič Brence, Branko Bregar, Katarina Vatovec, Tjaša Bertole, Marjeta Ferlan Istinič, Suzana Oreški, Matej Vinko

**Affiliations:** National Institute of Public Health, Trubarjeva cesta 2, 1000 Ljubljana, Slovenia; Angela Boškin Faculty of Health Care, Spodnji Plavž 3, 4270 Jesenice, Slovenia; Faculty of Medicine, University of Ljubljana, Vrazov trg 2, 1000 Ljubljana, Slovenia

## Abstract

**Introduction:** Frailty is a dynamic condition associated with increased vulnerability to adverse health outcomes in older adults. While previous research has primarily focused on deficit-based mental health factors, such as depression and loneliness, less is known about the role of positive mental health determinants, including well-being, resilience and social connectedness, in the development and progression of frailty. Understanding both risk and protective factors is essential for informing public health strategies aimed at promoting healthy ageing. This study aims to examine the longitudinal relationship between mental health and frailty in a nationally sampled population of adults aged 50 years and older in Slovenia.

**Methods and analysis:** This longitudinal observational study will collect data at four time points over a two-year period (January 2026-March 2028). A stratified random sample of community-dwelling adults aged 50–84 years will be drawn from the national population registry, with 5,000 individuals invited to participate in the first wave. Frailty, mental health and a set of social, psychological, and health-related factors will be assessed. Data will be analyzed using a combination of descriptive, inferential and longitudinal statistical methods to examine associations between frailty and mental health over time. Potential explanatory factors will also be explored within the longitudinal framework, and additional analyses will assess the impact of attrition.

**Ethics and dissemination:** The study has been approved by the Ethics and Deontology Committee of the National Institute of Public Health. Participation is voluntary, and informed consent will be obtained from all participants. Data will be anonymized and handled in accordance with applicable data protection regulations. Findings will be disseminated through peer-reviewed publications, conference presentations and public health reports to inform strategies for promoting healthy ageing.

**STRENGTHS AND LIMITATIONS OF THIS STUDY:**

- The study provides a multidimensional longitudinal assessment of frailty, incorporating both risk and protective factors.
- The four-wave design enables examination of bidirectional associations and within-person changes over time.
- Findings have direct public health relevance, informing prevention strategies by identifying modifiable psychological, health, and social determinants of frailty.
- The relatively short follow-up period (approximately two years) may limit conclusions about longer-term changes in frailty; however, prior research suggests that meaningful changes in frailty can occur within comparable time intervals.
- Attrition and selective non-response over time may affect the representativeness of the longitudinal sample.

## INTRODUCTION

Frailty is a dynamic state or process characterized by functional decline and reduced physiological reserves of an individual, which can lead to adverse health outcomes [1]. While the biopsychosocial model suggests that psychological and social factors contribute to this state, current research often fails to distinguish these as distinct longitudinal predictors of functional decline [2]. By framing frailty as a discrete clinical outcome, we can precisely evaluate how mental health trajectories—both deficit-and asset-based—drive or mitigate the progression of physiological vulnerability in older adults.

While traditional psychiatric epidemiology focuses on deficit-based models—primarily identifying depression and loneliness as drivers of functional decline—this approach overlooks the potential protective role of psychological assets [3,4]. Current evidence robustly links mental disorders to frailty risk, yet the longitudinal impact of positive mental health determinants, such as resilience and social inclusion, remains largely unquantified. Investigating these asset-based factors is essential for developing equitable public health interventions that move beyond symptom management toward health promotion in aging populations.

### Aims

This study aims to elucidate the complex interplay between mental health and frailty to inform evidence-based public health interventions centered on social equity. Specifically, we will characterize the associations between these constructs in Slovenians aged 50 and older, determine the longitudinal directionality of these links, and investigate potential psychological, social, and health mechanisms underlying these associations.

## METHODS AND ANALYSIS

### Study design

This longitudinal observational study will examine frailty and mental health, together with associated health, social and psychological variables. Repeated measurements will be obtained at four time points over a 27-month period (approximately 8 months apart). The study design and timing of measurements are illustrated in Figure 1. The study duration was selected to align with the project timeframe and is supported by evidence that frailty status is dynamic in community-dwelling adults, with meaningful transitions observed within one to two years[5].

**Figure 1.**
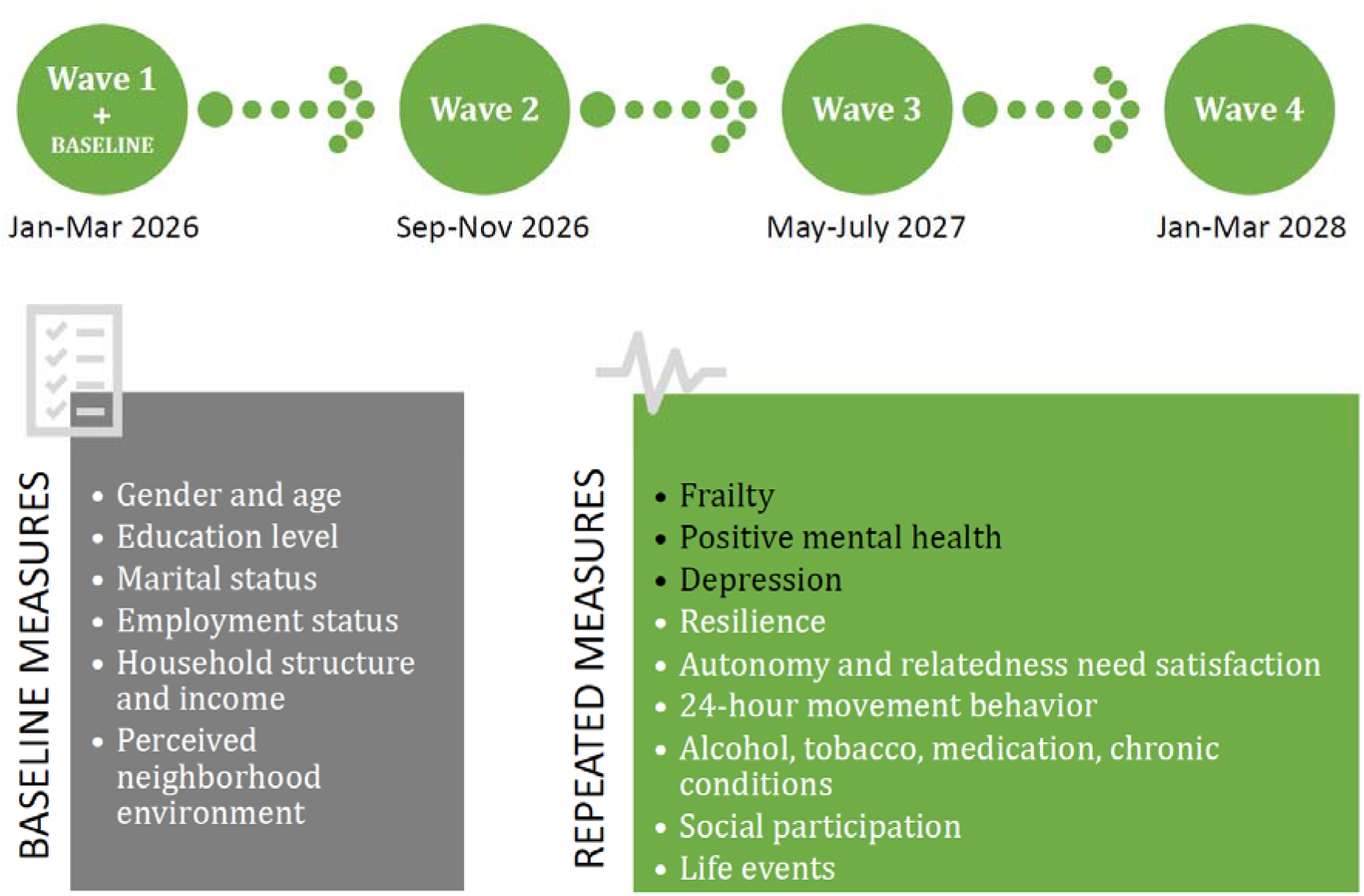
Study design and measurement timeline.

With at least three waves, it is possible to examine reciprocal associations between well-being and frailty as well as potential mechanisms, such as mediation, with appropriate temporal ordering[6]. The inclusion of a fourth wave strengthens the design by allowing assessment of lagged effects across multiple intervals, providing internal replication and improving robustness to attrition.

### Study setting

The study will be conducted in the Republic of Slovenia using a nationally sampled population of community-dwelling adults aged 50–84 years.

### Participants and sampling

The target population involves community-dwelling residents of the Republic of Slovenia aged 50–84 years. The lower age limit was selected to capture early frailty processes beginning in midlife [7], while the upper limit reduces attrition associated with advanced age while retaining individuals at high risk of frailty progression. Individuals residing in institutional care settings (e.g., nursing homes or other long-term care institutions) will be excluded, as frailty prevalence and associated health characteristics differ substantially between institutionalized and community-dwelling populations [8].

The sample will be prepared by the Statistical Office of the Republic of Slovenia (SURS) using a stratified simple random sampling design. Stratification by statistical region, settlement type, and age group ensures geographic representativeness and enables subgroup comparisons. Approximately equal representation of the three age groups (50–64, 65–74, 75–84 years) is planned to facilitate comparisons across age categories.

### Measures

#### Primary measures

Frailty will be assessed using the Tilburg Frailty Indicator (TFI), a 15-item instrument covering physical, psychological and social domains [9]. The TFI has demonstrated good reliability and validity, including in community-dwelling older adults in Slovenia [8].

Positive mental health will be measured using the Mental Health Continuum – Short Form (MHC-SF), a 14-item scale assessing emotional, social and psychological well-being [10]. The instrument has shown good psychometric properties in international studies [10–12].

Depressive symptoms will be assessed using the Patient Health Questionnaire–9 (PHQ-9), a widely used measure of depressive symptom severity [13]. The PHQ-9 has demonstrated good reliability and validity, including in Slovenian samples [14].

Additional details on all measures, including item examples and psychometric properties, are provided in Supplementary Material.

Psychological, health-related, and social factors Resilience will be measured using the 10-item Connor–Davidson Resilience Scale (CD-RISC-10), which assesses adaptability, self-efficacy, emotional regulation, optimism, and cognitive focus [15]. The scale has demonstrated good psychometric properties in Slovenian adults [16].

Basic psychological need satisfaction of autonomy and relatedness will be assessed using selected subscales from the Basic Psychological Need Satisfaction and Frustration Scale (BPNSFS) [17]. The instrument has been shown to be reliable and valid across different cultural contexts [18,19].

Physical activity, sedentary behavior and sleep (i.e. 24-hour movement behavior) will be assessed using the GIB24 questionnaire, a 10-item instrument developed and validated in Slovenia [20].

Additional health-related factors, including alcohol use, tobacco use, medication use, and chronic conditions, will also be assessed using study-specific items.

Social participation will be assessed through involvement in community and voluntary organizations (e.g., cultural, religious or hobby-based groups), as well as frequency of engagement in leisure-time activities.

Recent positive and negative life events will be assessed using study-specific items designed to capture relevant contextual factors.

#### Baseline measures

Participants will provide information on sociodemographic characteristics, including gender, year of birth, marital status, number of household members, employment status, education level, and household income.

Perceived neighborhood environment (e.g., access to services, green spaces, safety and environmental quality) will be assessed at baseline using study-specific items.

#### Timing of measurements

Primary variables, as well as psychological, social, and health-related factors will be assessed at each wave of data collection. These measures refer to recent time periods (e.g., the past month or past 6 months), enabling the analysis of within-person changes over time. Sociodemographic characteristics will be collected at baseline and treated as time-invariant variables in the analyses.

### Recruitment and data collection procedures

In the first wave, all individuals in the sample will receive a postal invitation package containing a paper questionnaire and an information letter. Participants will have the option to complete the questionnaire either in paper form and return it by mail or electronically via an online version accessible through a QR code printed on the questionnaire.

Participants will be informed at the outset that the study consists of four waves and that they will be invited to participate in subsequent waves. In the first wave, participants will be offered the opportunity to voluntarily provide their email address or telephone number if they wish to be contacted electronically or by phone for future waves.

Participation is voluntary and may be discontinued at any time without consequences. Completion of the questionnaire will be considered as provision of informed consent.

The estimated time required to complete the questionnaire is approximately 20–25 minutes, based on pilot testing conducted with a small group of participants. To acknowledge participants’ time and effort, those who participate in at least three waves will receive a small symbolic gift in the form of project promotional material. Information about this incentive is clearly stated in the initial invitation.

### Statistical analysis

Descriptive statistics will summarize sample characteristics and study variables at each wave. Differences between respondents and non-respondents will be examined to assess potential selection bias. Group differences in categorical variables will be assessed using χ^2^ tests, and differences in continuous variables using t-tests or non-parametric alternatives, as appropriate.

Frailty and mental health will be analyzed using validated multi-item instruments. Where appropriate, latent variable modelling approaches will be applied to account for measurement properties. Confirmatory factor analyses will be conducted to evaluate the measurement properties of the constructs across waves, and model fit will be assessed using standard indices such as the Comparative Fit Index (CFI), Tucker–Lewis Index (TLI), Root Mean Square Error of Approximation (RMSEA), and Standardized Root Mean Square Residual (SRMR).

Longitudinal associations between frailty and mental health will be examined using structural equation modelling or related longitudinal modelling approaches for panel data across four waves. Cross-lagged panel models, including random-intercept extensions [21], will be used to examine temporal associations while accounting for both between-person differences and within-person changes over time.

Health-related, social and psychological variables will then be examined as potential explanatory factors in focused longitudinal analyses. Baseline sociodemographic characteristics and other time-invariant variables will be included as covariates in the longitudinal models.

Missing data will be handled using appropriate statistical methods depending on the extent and pattern of missingness. In longitudinal models, approaches that allow the use of all available data will be applied where appropriate. Sensitivity analyses will be conducted using alternative model specifications, including traditional cross-lagged panel models, to assess the robustness of findings.

### Sample size and statistical power

A Monte Carlo simulation was conducted in Mplus 8.8 to evaluate statistical power for the planned longitudinal panel analyses, using a random-intercept cross-lagged panel model (RI-CLPM) as the primary reference model with four waves of data and 85% retention per wave. The simulation assumed autoregressive effects of 0.50 to 0.60, reflecting moderate temporal stability and consistent with longitudinal cross-lagged analyses reporting similar stability for frailty-related and psychological constructs [22], as well as cross-lagged effects of absolute magnitude 0.07 between frailty and mental health. This value corresponds to a moderate cross-lagged effect according to recent empirical benchmarks for cross-lagged models [23].

With an initial sample size of 1,500 participants and 2,000 Monte Carlo replications, the simulation indicated adequate power to detect the hypothesized cross-lagged effects, with power of approximately 0.83 for the pathway from frailty to subsequent mental health and 0.80 for the pathway from mental health to subsequent frailty. Results suggest that a baseline sample of approximately 1,500 participants, assuming an average retention rate of 85% per wave across four repeated measurements, is sufficient for the planned longitudinal analyses.

To achieve the required baseline sample, 5,000 individuals will be invited to participate in wave 1 from the sampling frame prepared by SURS. Recent national health surveys conducted by the National Institute of Public Health in Slovenia (NIJZ) have reported response rates of approximately 41% to 67%. However, these surveys were cross-sectional and included broader adult age groups. Because the present study targets older adults and includes four waves of data collection, cumulative attrition is expected. Therefore, a conservative recruitment target was chosen to ensure a sufficient baseline sample for the planned longitudinal analyses.

### Attrition and mortality linkage

Additional analyses will compare participants who remain in the study with those who drop out in subsequent waves. To assess potential reasons for attrition in follow-up waves, a one-time secure linkage will be conducted between study data and the official mortality database maintained by the NIJZ. The purpose of this linkage is to determine whether non-participation in follow-up waves is attributable to participant death.

For participants who complete at least one wave and subsequently discontinue participation, information on mortality is analytically relevant because death represents a competing event that may occur prior to the development of frailty or changes in mental health. Death will therefore be treated differently from other forms of missing data due to attrition. The linkage will be conducted in accordance with applicable data protection regulations and secure data handling procedures.

### Patient and public involvement

Patients and the public were not directly involved in the design or planned conduct of this study and will not be directly involved in the reporting or dissemination of the findings. This study is based on a nationally representative population sample and is not focused on a specific patient group; therefore, no single patient or public group was considered appropriate to involve. The primary target audience includes researchers and stakeholders in public mental health and ageing research.

## Data Availability

No datasets were generated or analysed for this study protocol. Data will be collected as part of the study and will be available from the corresponding author upon reasonable request, in accordance with applicable data protection regulations.

## ETHICS AND DISSEMINATION

This study is supported by an operation funded under the European Cohesion Policy Programme 2021–2027 in Slovenia, Priority 7: “Long-term Care and Health, and Social Inclusion”, Specific Objective ESF+ 4.11. The operation is co-financed by the European Union through the European Social Fund Plus (ESF+) and by the Republic of Slovenia from the national budget and is implemented as part of the project “Systemic Approach to Frailty with an Emphasis on Prefrailty for Healthy and Quality Ageing.” The funders had no role in the design of the study protocol and will have no role in the collection, analysis, or interpretation of data, nor in the writing of the manuscript or the decision to publish the results.

The study has been approved by the “Ethics and Deontology Committee of the National Institute of Public Health” (No. 631-2/2025.29 (013), 6 February 2026). In the first wave of data collection, participants will receive an information letter containing detailed information about the study, including its purpose, procedures, voluntary participation, and measures taken to ensure confidentiality. Participants will be informed that completion of the questionnaire implies informed consent. Participation is voluntary, and participants may withdraw at any time without consequences.

Findings will be disseminated through peer-reviewed open access publications and presentations at national and international public health conferences.

## Authors’ contributions

MV and PMB conceived the study idea. PMB, MV, BB, MFI, SO and TB contributed to the study and study protocol design. MV, PMB, BB, and KV contributed to writing and refining the manuscript. All authors critically reviewed and approved the final version of the manuscript.

