## Supplementary Material for "Study protocol: Longitudinal observational study on frailty and mental health"

**Supplementary Table 1** Detailed description of study measures.

| **Measure** | **Instrument** | **Author(s), year** | **Example item** | **Psychometric properties** |
| --- | --- | --- | --- | --- |
| **Baseline measures** | | | | |
| Sociodemographic characteristics | 8 standard sociodemographic items: gender, year of birth, marital status, number of adult and minor household members, employment status, education level, and household income | / | What is the highest level of education you have completed? | / |
| Perceived neighborhood environment | 8 items regarding access to services, green spaces, safety and environmental quality | / | Do you have access to green and other recreational areas, as well as the most important services, within walking distance, by bicycle, or by public transport in your area? | / |
| **Primary repeated measures** | | | | |
| Frailty | Tilburg Frailty Indicator (TFI) | Gobbens et al., 2010 | Do you feel physically healthy? | Cronbach’s α = 0.79, ICC = 0.79 |
| Positive mental health | Mental Health Continuum – Short Form (MHC-SF) | Keyes et al., 2008 | During the past month, how often did you feel happy? | Cronbach’s α = .89, confirmatory factor analysis supports construct validity |
| Depressive symptoms | Patient Health Questionnaire–9 (PHQ-9) | Spitzer, 1999 | Over the last 2 weeks, how often have you been bothered by any of the following problems? Feeling down, depressed, or hopeless. | Cronbach’s α = 0.87, AUC = 0.92 |
| **Psychological, health-related, and social repeated measures** | | | | |
| Resilience | Connor–Davidson Resilience Scale (CD-RISC-10) | Campbell‐Sills & Stein, 2007 | I am able to adapt when changes occur. | Cronbach’s α > .76, convergent validity supported with related constructs |
| Autonomy satisfaction and relatedness satisfaction | Basic Psychological Need Satisfaction and Frustration Scale (BPNSFS) | Chen et al., 2015 | I feel a sense of choice and freedom in the things I undertake. | Cronbach’s α = .80, validity supported by related constructs |
| **Measure** | **Instrument** | **Author(s), year** | **Example item** | **Psychometric properties** |
| Physical activity, sedentary behavior and sleep (i.e. 24-hour movement behavior) | GIB24 | Kastelic et al., 2025 | On how many days in a typical week did you engage in household activities at or around your home that made your breathing faster? | ICC = 0.51-0.71, validity supported by comparison to established reference instruments |
| Chronic conditions | 7 items assessing different health conditions and the timing of diagnosis (within the past 6 months vs. earlier) | / | Do you have any of the following diseases or conditions that have been diagnosed by a doctor? Cancer | / |
| Alcohol use | 4 items assessing the amount and frequency of alcohol consumption | / | How often have you consumed drinks containing alcohol in the past 6 months? | / |
| Tobacco use | 1 item regarding current or past tobacco use | / | Do you smoke tobacco (cigarettes, cigars, cigarillos, or pipe tobacco)? | / |
| Medication and dietary supplement use | 2 items assessing the consumption of medication and dietary supplements | / | How many different medications prescribed by your doctor are you currently taking? | / |
| Recent negative life events | 8 items assessing the occurrence of negative life events in the last 6 months | / | Have you experienced the following event in the past 6 months: death of a loved one? | / |
| Recent positive life events | 6 items assessing the occurrence of positive life events in the last 6 months | / | Have you experienced the following event in the past 6 months: a significant improvement in your own health? | / |
| Social participation | 1 item assessing frequency and 1 item assessing type of social participation (8 response options, e.g., religious groups, cultural and artistic associations) | / | Approximately how many times in the past month have you participated in leisure activities (e.g., cinema, restaurants, sports, cultural events)? | / |

*Note:* Measures of perceived neighborhood environment, additional health-related factors: chronic conditions, alcohol use, tobacco use, medication and dietary supplement use, recent negative life events, recent positive life events, and social participation were developed by the research team for the purposes of this study or adapted from standard questions used in national surveys conducted by the National Institute of Public Health (NIJZ).
